# Implementation of Rapid Genome Sequencing for Infants with Congenital Heart Disease

**DOI:** 10.1101/2022.12.16.22283479

**Authors:** Thomas Hays, Rebecca Hernan, Michele Disco, Emily Griffin, Nimrod Goldshtrom, Diana Vargas, Ganga Krishnamurthy, Atteeq U. Rehman, Amanda T. Wilson, Saurav Guha, Shruti Phadke, Volkan Okur, Dino Robinson, Vanessa Felice, Avinash Abhyankar, Vaidehi Jobanputra, Wendy K. Chung

## Abstract

**Background:** Rapid genome sequencing (rGS) has been shown to improve the care of critically ill infants. Congenital heart disease (CHD) is a leading cause of infant mortality, and is often caused by genetic disorders, yet the utility of rGS has not been prospectively studied in this population.

**Methods:** We conducted a prospective evaluation of the use of rGS to improve the care of infants with CHD in our cardiac neonatal intensive care unit (CNICU).

**Results:** In a cohort of 48 infants with CHD, rGS diagnosed 14 genetic disorders in 13 (27%) individuals and led to changes in clinical management in eight (62%) cases with diagnostic results. These included two cases in whom genetic diagnoses helped avert intensive, futile interventions prior to CNICU discharge, as well as three cases in whom eye disease was diagnosed and treated in early childhood. Genetic disorders were associated with small for gestational age birth weight.

**Conclusions:** Our study provides the first prospective evaluation of rGS for infants with CHD to our knowledge. We found that rGS diagnosed genetic disorders in 27% of cases and led to changes in management in 62% of cases with diagnostic results. Our model of care was enabled by multidisciplinary coordination between neonatologists, cardiologists, surgeons, geneticists, and genetic counselors. These findings highlight the important role for rGS in CHD and demonstrate the need for expanded study of how to implement this resource to a broader population of infants with CHD.

## Introduction

Recent studies using genomic sequencing have demonstrated that genetic disorders account for a significant portion of critical illness in infants.^1-5^ Rapid genome sequencing (rGS) leads to improvements in clinical management and is perceived as beneficial by healthcare providers and parents of critically ill infants.^6-11^ Genetic disorders in infants often present nonspecifically, then progress to life threatening illness or death.^12,13^ For example, it may be impossible to distinguish whether a congenital anomaly is an isolated feature, or the sentinel manifestation of a syndromic process that includes progressive multi-organ disease. Thus, a rapid diagnosis can help prevent or mitigate disease processes, avoid aggressive but futile interventions, and shorten the diagnostic odyssey.

Congenital heart disease (CHD), which affects approximately 1% of infants, is often genetic in etiology.^14-16^ Pathogenic copy number variants (CNVs) and single and oligonucleotide variants can be found in approximately 10–20% of individuals with CHD, with increasingly recognized contribution from noncoding variants.^17^ Genetic disorders are particularly prevalent in infants with complex structural anomalies and involvement of other organ systems.^18-20^ Infants with CHD are at high risk for severe co-morbidities and mortality.^21^ Retrospective analysis of rGS in infants with CHD demonstrated improved care at a lower cost;^22^ however, rGS has not been prospectively studied in this population. Here we describe a prospective investigation of the use of rGS to improve the care of infants with CHD.

## Methods

This was a single-center prospective cohort study, which was approved by the Columbia University Irving Medical Center Institutional Review Board. Parents of infants gave written informed consent, and procedures were followed in accordance with institutional guidelines. We implemented a novel, multidisciplinary strategy to integrate rGS into the Columbia model of neonatal cardiac care.^23,24^ In this model, infants with CHD are cared for in a dedicated cardiac neonatal intensive care unit (CNICU), staffed by medical and nursing providers with specialized training and expertise caring for infants with CHD. To decrease the time to return of results, neonatologists and cardiologists were empowered to directly identify infants in whom: 1) a genetic disorder was suspected, and 2) rapid diagnosis might improve clinical management. Cases were then directly referred to a team of genetic counselors to confirm appropriateness of rGS testing, and to conduct pretest counseling. This counseling included discussion of the return of primary genetic results, secondary medically actionable findings, and selected pharmacogenomic variants with neonatal management implications. Genetic counselors and geneticists performed post-test counseling. Infants with any structural CHD without an established genetic diagnosis, and with both parents available for sample collection for trio genome sequencing were eligible for study inclusion.

rGS was performed in a Clinical Laboratory Improvement Amendments (CLIA) approved laboratory. Total genomic DNA was purified from whole blood using standard methods. Sequencing libraries were then prepared using the KAPA Hyper Library preparation kit (Kapa Biosystems, Wilmington, MA). Libraries were subjected to 2 X 150 bp paired-end sequencing on HiSeq X instrument (Illumina, USA), to achieve a mean sequencing depth of 30 X. Variant analysis was performed using a proprietary clinical pipeline at the New York Genome Center’s clinical laboratory which follows the GATK best practices guidelines.^25^ Variant interpretation and classifications were performed using standard criteria of the American College of Medical Genetics and Genomics (ACMG).^26-28^ Results of rGS were considered diagnostic for pathogenic or likely pathogenic variants in genes with consistent phenotypes and patterns of inheritance as observed.

A complete manual chart review was conducted from admission to discharge from the CNICU. Demographic and clinical details were collected including gestational age, birth weight, the presence of extracardiac anomalies, incidence of co-morbidities and mortality, family history, and disposition from the CNICU. Birth weight was classified as appropriate, small, or large for gestational age (AGA, SGA, or LGA) based on the 10^th^ and 90^th^ percentiles for gestational age and sex by Fenton classification.^29^ Forms of CHD were hierarchically categorized into groups with similar etiology, as previously described.^30^ First, cases involving a laterality defect (LAT), including heterotaxy, and regardless of the presence of other lesions, were grouped. Next, cases with a conotruncal defect (CTD) were grouped regardless of the presence of other defects. Then, cases with an atrioventricular septum defect (AVCD) with normally related great vessels were grouped. Then, cases involving left or right ventricular outflow tract obstruction (LVOT or RVOT) with normally related great vessels were grouped. The remaining cases were grouped together and consisted of infants with an isolated atrial septum defect or a pulmonary vein anomaly (ASPV). To test which characteristics were associated with diagnostic rGS results, multiple variable logistic regression was performed. Infants born AGA and LGA were grouped together because all four LGA infants had nondiagnostic rGS results. CHD categorization was excluded from the regression because no diagnostic rGS results were found in infants with AVCD and RVOT anomalies. Family history was similarly excluded as no individuals with diagnostic results had a known family history of CHD. Analyses were made using RStudio (version 2022.02.0).^31^ Author TH had full access to all data in the study and takes responsibility for its integrity and the data analysis.

## Results

Individuals were recruited from June 2020 through March 2022. The median (interquartile range (IQR)) age at testing was 7.5 (5.3) days. The cohort consisted of 48 infants (Table 1) including 17 females and 31 males. No significant differences were observed in baseline characteristics, except in birth weight for gestational age (P = 0.003 by chi-square test). Six (46.2%) infants with diagnostic rGS results were born SGA, while only two (5.7%) infants with nondiagnostic rGS infants were born SGA. The median (IQR) gestational age was 38.0 (2.0) weeks, and the median (IQR) birth weight was 2857.5 (785.0) g. Extracardiac anomalies were present in 38 (79.2%) infants. Cardiac lesions were categorized as ASPV (n = 4 (8.3%)), AVCD (n = 2 (4.2%)), CTD (n = 20 (41.7%)), LAT (n = 7 (14.6%)), LVOT (n = 13 (27.1%)), and RVOT (n = 2 (4.2%)) (Table 2). In five cases (10.4%) there was a family history of CHD. Seven infants (14.6%) died prior to CNICU discharge, 24 (50.0%) were discharged home, nine (18.8%) were transferred to rehabilitation facilities, and eight (16.7%) were transferred to other institutions following surgery and postoperative care.

**Table 1.** Characteristics of 48 infants with congenital heart disease.

| Characteristic | Nondiagnostic<br>rGS<br>(N=35) | Diagnostic rGS<br>(N=13) | P-value | Diagnostic<br>Yield of rGS |
| --- | --- | --- | --- | --- |
| Sex |  |  |  |  |
| Female | 12 (34.3%) | 5 (38.5%) | 1.000 | 0.294 |
| Male | 23 (65.7%) | 8 (61.5%) |  | 0.256 |
| Gestational Age<br>(weeks) |  |  |  |  |
| Mean (SD) | 37.9 (2.13) | 38.5 (1.51) | 0.304 | NA |
| Median [Min,<br>Max] | 38.0 [30.0, 41.0] | 39.0 [36.0, 41.0] |  |  |
| Birth Weight (g) |  |  |  |  |
| Mean (SD) | 2980 (689) | 2680 (669) | 0.177 | NA |
| Median [Min,<br>Max] | 3070 [1260, 4390] | 2840 [1530, 3840] |  |  |
| Birth Weight for<br>Gestational Age |  |  |  |  |
| AGA | 29 (82.9%) | 7 (53.8%) | 0.003 | NA |
| LGA | 4 (11.4%) | 0 (0%) |  |  |
| SGA | 2 (5.7%) | 6 (46.2%) |  |  |
| Cardiac Anomaly |  |  |  |  |
| ASPV | 2 (5.7%) | 2 (15.4%) | 0.561 | 0.500 |
| AVCD | 2 (5.7%) | 0 (0%) |  | 0.000 |
| CTD | 16 (45.7%) | 4 (30.8%) |  | 0.200 |
| LAT | 5 (14.3%) | 2 (15.4%) |  | 0.286 |
| LVOT | 8 (22.9%) | 5 (38.5%) |  | 0.385 |
| RVOT | 4 (11.4%) | 0 (0%) |  | 0.000 |
| Extracardiac<br>Anomaly |  |  |  |  |
| Absent | 8 (22.9%) | 2 (15.4%) | 0.868 | 0.200 |
| Present | 27 (77.1%) | 11 (84.6%) |  | 0.289 |
| Known Family<br>History of CHD |  |  |  |  |
| Absent or none<br>known | 30 (85.7%) | 13 (100%) | 0.364 | 0.302 |
| Present | 5 (14.3%) | 0 (0%) |  | 0.000 |

| Characteristic | Nondiagnostic rGS (N=35) | Diagnostic rGS (N=13) | P-value | Diagnostic Yield of rGS |
| --- | --- | --- | --- | --- |
| **Disposition** |  |  |  |  |
| Death | 5 (14.3%) | 2 (15.4%) | 0.895 | 0.286 |
| Home | 18 (51.4%) | 6 (46.2%) |  | 0.250 |
| Rehabilitation Facility | 7 (20.0%) | 2 (15.4%) |  | 0.222 |
| Transfer | 5 (14.3%) | 3 (23.1%) |  | 0.375 |

P-value calculated by chi-square or t-test as indicated for each characteristic.
Diagnostic yield is defined as the rate of diagnostic rGS within individuals in a row.
Abbreviations: AGA (appropriate for gestational age), ASPV (atrial septal defect or pulmonary vein anomaly), AVCD (atrioventricular septal defect), CHD (congenital heart disease), CTD (conotruncal heart defect), LAT (laterality disorder), LVOT (left ventricular outflow tract obstruction), OMIM (Online Mendelian Inheritance in Man), rGS (rapid genome sequencing), RVOT (right ventricular outflow tract obstruction), SGA (small for gestational age).

**Table 2.** Cardiac anomalies in 48 infants.

| **Cardiac Anomaly** | **Number (%)** |
| --- | --- |
| **Atrial Septal Defect or Pulmonary Vein Anomaly** | **4 (8.3%)** |
| Atrial septal defect and biventricular dysfunction | 1 (2.1%) |
| Pulmonary vein stenosis | 1 (2.1%) |
| Total anomalous pulmonary venous connection | 2 (4.2%) |
| **Atrioventricular Septal Defect** | **2 (4.2%)** |
| Atrioventricular septal defect | 2 (4.2%) |
| **Conotruncal Defect** | **20 (41.7%)** |
| Aortopulmonary window | 1 (2.1%) |
| D-transposition of the great arteries | 3 (6.3%) |
| Double inlet left ventricle with L-transposition of the great arteries | 3 (6.3%) |
| Double outlet right ventricle | 4 (8.3%) |
| Interrupted aortic arch | 1 (2.1%) |
| Truncus arteriosus | 2 (4.2%) |
| Tetralogy of Fallot | 6 (12.5%) |
| **Laterality Defect** | **7 (14.6%)** |
| Dextrocardia with hypoplastic left heart and total anomalous pulmonary venous connection | 1 (2.1%) |
| Heterotaxy with double outlet right ventricle | 1 (2.1%) |
| Heterotaxy with double outlet right ventricle and total anomalous pulmonary venous connection | 1 (2.1%) |
| Heterotaxy with hypoplastic left heart, double outlet right ventricle, and total anomalous pulmonary venous connection | 1 (2.1%) |
| Heterotaxy with L-transposition and total anomalous pulmonary venous connection | 1 (2.1%) |
| Heterotaxy with pulmonary atresia and partial anomalous pulmonary venous connection | 1 (2.1%) |
| Heterotaxy with atrioventricular septal defect and coarctation of the aorta | 1 (2.1%) |
| **Left Ventricular Outflow Tract Obstruction** | **13 (27.1%)** |
| Coarctation of the aorta | 6 (12.5%) |
| Coarctation of the aorta and total anomalous pulmonary venous connection | 2 (4.2%) |
| Hypoplastic left heart | 5 (10.4%) |
| **Right Ventricular Outflow Tract Obstruction** | **2 (4.2%)** |
| Pulmonary atresia | 2 (4.2%) |

Genetic disorders were diagnosed in 13 (27.1%) infants, 12 of which related to CHD and one which was a secondary diagnosis (Table 3). These diagnoses consisted of nine single gene disorders four CNVs, including three diagnoses of cat eye syndrome. One infant had dual CNV diagnoses. Of the 14 distinct genetic disorders, all but two were *de novo* variants. The secondary diagnosis was autosomal dominant polycystic kidney disease in a family with a known history of polycystic kidney disease. This individual was also found to have a *de novo* variant of uncertain significance in *SMAD6*, which might contribute to the cardiac phenotype and facial dysmorphisms. No significant differences were found in the baseline characteristics of infants with diagnostic as compared to those with nondiagnostic rGS results, except for birth weight for gestational age (Table 1). SGA birth occurred in six (46.2%) infants with diagnostic rGS, as compared to two (5.7%) with nondiagnostic results. LGA birth was not found in infants with diagnostic rGS, whereas it occurred in four (11.4%) of infants with nondiagnostic rGS. Multiple variable logistic regression was performed to test whether any individual characteristics were associated with diagnostic rGS results when controlling for other characteristics. This demonstrated that SGA birth was strongly associated with the presence of a genetic disorder (adjusted OR of 93.77 (95% CI = 6.60, 7684.49); P-Value 0.007) (Figure 1).

**Table 3.** Cases with Diagnostic rGS results.

| Study ID | Gene | Variant, Zygosity, Inheritance, Classification | Disease Association (Inheritance, OMIM number) | Cardiac Anomaly | Extracardiac Features | Disposition | Change in Management |
| --- | --- | --- | --- | --- | --- | --- | --- |
| 91007 | *PTPN11* | NM_002834.4: c.179G>C (p.Gly60Ala), heterozygous, *de novo*, pathogenic | Noonan syndrome 1 (AD, 163950); leopard syndrome (AD, 151100); metachondromatosis (AD, 156250) | heterotaxy, HLHS, DORV, TAPVC | craniosynostosis, external ear malformation, ventriculomegaly | death following redirection of care with presumed sepsis and multiorgan failure | NC |
| 91014 | *FOXJ1* | NM_001454.4: c.867dup (p.Ser290Glnfs Ter12), heterozygous, *de novo*, pathogenic | ciliary dyskinesia, primary, 43 (AD, 618699) | heterotaxy, PA, TAPVC | fetal growth restriction | transfer | referral to pulmonology; surveillance for hydrocephalus |
| 91016 | *PUF60* | NM_078480.2: c.24+1G>A, heterozygous, *de novo*, likely pathogenic | Verheij syndrome (AD, 615583) | DORV | refractive error (bilateral hyperopia and anisometropia), renal duplication | home | referral to ophthalmology identified refractive error to be corrected with glasses |
| 91020 | FOXF1 | NM_001451.3: c.691_698del (p.Ala231Argfs Ter61), heterozygous, *de novo*, pathogenic | alveolar capillary dysplasia with misalignment of pulmonary veins (AD, 265380) | VSD, CoA | omphalocele, respiratory failure | death secondary to respiratory failure | change in ECMO and surgical candidacy |
| 91025 | MYH6 | NM_002471.3: c.1138G>A (p.Glu380Lys), heterozygous, *de novo*, likely pathogenic | sick sinus syndrome 3 (614089); cardiomyopathy, delated 1EE (AD, 613252); cardiomyopathy, hypertrophic, 14 (AD, 613251); atrial septal defect 3 (614090) | DORV | pentalogy of Cantrell | rehabilitation facility | surveillance for heart failure |
| 91026 | - | gain (mosaic), seq[GRCh38]2 2q11.1–q11.2g.(?_167 63316)_ (21656 291_?)dup, de novo, pathogenic<br>gain (mosaic), seq[GRCh38]2 2q13.31–q13.33(?_4477 2490_5067130 7_?) dup, de novo, likely pathogenic<br>loss, seq[GRCh38]2 2q13.33g. (50671952_50 781160)del, de novo, pathogenic | cat eye syndrome (AD, 115470) | CoA, TAPVC | fetal growth restriction, refractive error (bilateral hyperopia), strabismus (unilateral nonaccommodative esotropia), solitary kidney with reflux, dysmorphic features, absent ductus venosus | home | Referral to ophthalmology identified refractive error and strabismus managed with corrective glasses and patching |
| 91029 | - | gain seq[GRCh38]2 2q11.21g. (?_16568030)_ (18191862_?)d up, *de novo*, pathogenic | cat eye syndrome (AD, 115470) | TAPVC | congenital hypothyroidism | home | Referral to ophthalmology identified coloboma |
| 91030 | *KMT2D* | NM_003482.3: c.9880_9894de lins CCCTGCC (p.Ala3294Profs*4), heterozygous, *de novo*, pathogenic | kabuki syndrome 1 (AD, 147920) | HLHS (MS/AS) | microcephaly, dysmorphic features | transfer | redirection to comfort care goals; avoidance of cardiac surgery |
| 91034 | *MT-ATP6* | NC_012920.1: m.8969G>A (p.Ser148Asn), heteroplasmic (93% VAF), *de novo*, likely pathogenic | ATP-6 associated mitochondrial disorder (516060) | CoA and HOCM | PAH, bilateral hydronephrosis, epispadias, cryptorchidism, micropenis, dysplastic nails, thrombocytopenia, choroid plexus cyst | rehabilitation facility | referral to neurology; prescription of co-factors; preventive surveillance for metabolic disorder |
| 91035 | CHD7 | NM_017780.4: c.1879dup (p.Gln627Profs Ter5), heterozygous, *de novo*, pathogenic | CHARGE syndrome (AD, 214800); hypogonadotropic hypogonadism 5 with or without anosmia (AD, 612370) | ToF | Colobomas, left microphthalmia, cleft lip and palate, cryptorchidism, micropenis, undescended testes, intraventricular hemorrhage | transfer | NC |
| 91036 | - | gain, seq[GRCH38]2 2q11.21g. (?_16568033)_ (18199587_?)dup, *de novo*, pathogenic | cat eye syndrome (AD, 115470) | TAPVC | Colobomas affecting right iris, left lens, and bilateral retinas, refractive error (bilateral hyperopia), strabismus (Duane's syndrome), unilateral microphthalmia | home | referral to ophthalmology identified severe coloboma and risk of unilateral blindness; prescription of protective glasses |
| 91039 | - | gain,<br>seq[GRCh38]1<br>q21.1g.(14605<br>5940_1470077<br>08)_(14836667<br>6_<br>148925963)<br>dup,<br>*de novo*,<br>pathogenic<br>and<br>gain,<br>seq[GRCh38]1<br>6p13.11g.(146<br>79387_150326<br>10)_(16203929<br>_16530762)<br>dup,<br>*de novo*,<br>likely<br>pathogenic | 1q21.1<br>microduplication<br>syndrome (AD,<br>612474); and<br>16p13.11<br>microduplication<br>syndrome | IAA, AP<br>window | dysmorphic<br>features | home | NC |
| 91044 | PKD1 | NM_00100994<br>4.2:c.6795C>G<br>(p.Tyr2265Ter)<br>,<br>heterozygous,<br>paternal,<br>pathogenic | polycystic kidney<br>disease (AD,<br>173900) | CoA, ASD | dysmorphic<br>features | home | NC |

Abbreviations: AD (autosomal dominant), AP (aortopulmonary), AS (aortic stenosis), ASD (atrial septal defect), CoA (coarctation of the aorta), DORV (double outlet right ventricle), HLHS (hypoplastic left heart syndrome), HOCM (hypertrophic cardiomyopathy), IAA (interrupted aortic arch), LV (left ventricle), MS (mitral stenosis), NC (no change), PA (pulmonary atresia), PAH (pulmonary arterial hypertension), PS (pulmonary stenosis), TAPVC (total anomalous pulmonary venous connection), TGA (transposition of the great arteries), ToF (tetralogy of Fallot), VAF (variant allele frequency), VSD (ventricular septal defect).

**Figure 1.**
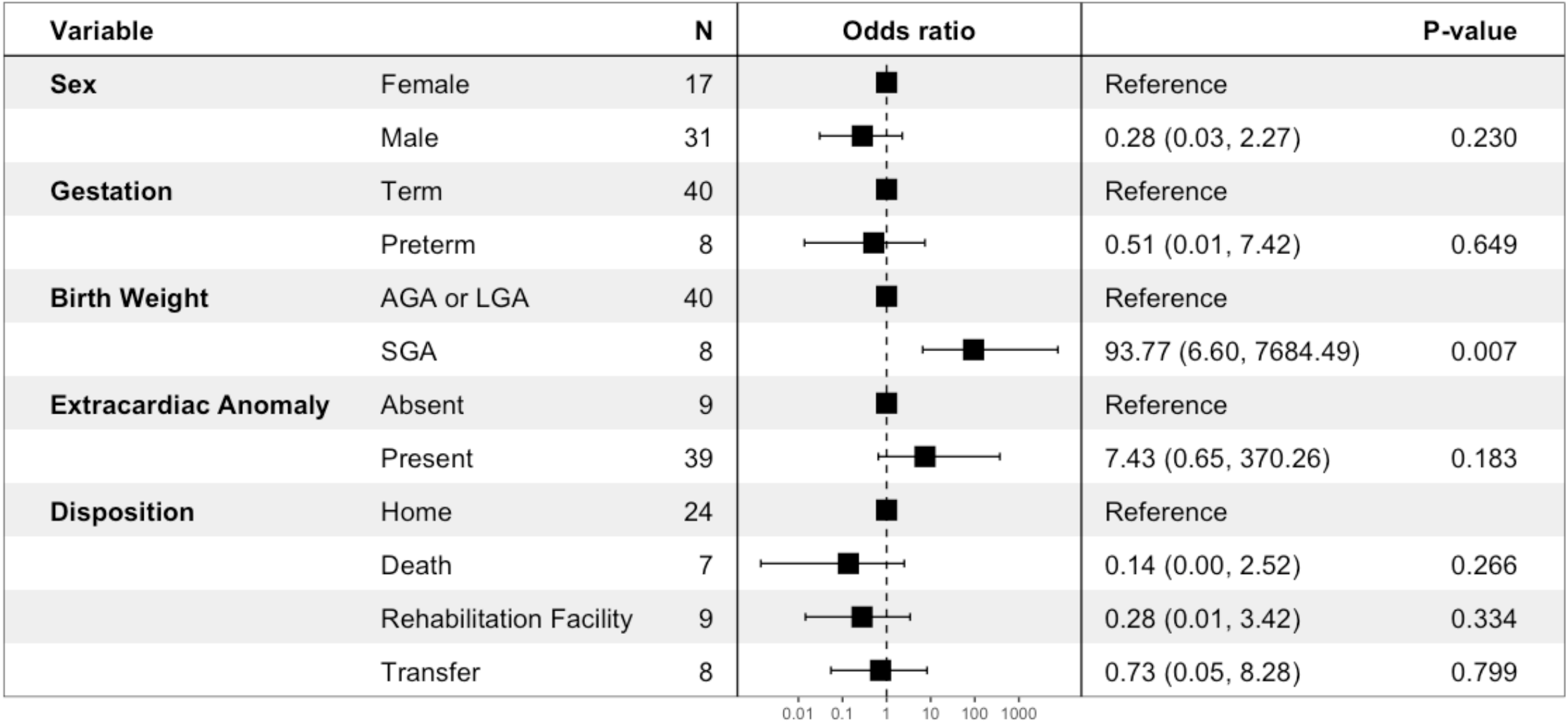
Multiple variable logistic regression for the association of characteristics with diagnostic rGS results. Input variables consisted of sex, gestation (term or preterm), birth weight (appropriate, small, or large for gestational age (AGA, LGA, or SGA) the presence of extracardiac anomalies, and disposition. The adjusted odds ratio (95% CI) and Bonferroni-corrected P-value are shown. Only SGA birth was found to have a significant association with diagnostic rGS results.

Management changes that resulted from rGS results occurred in eight of 13 (61.5%) cases. These changes included referral to subspecialists from other disciplines (ophthalmology, pulmonology, and neurology) for treatment and screening for manifestations associated with genetic disorders, tailoring of medical therapy, and surveillance for metabolic decompensation. The subspeciality service to which referrals were most often made was ophthalmology. The four infants referred for ophthalmologic evaluations were each found to have eye disease related to their genetic disorders, which led to interventions in three cases including corrective glasses for refractive error and patching for strabismus. In two cases, rGS results enabled the CNICU team to redirect goals of care and avoid intensive, futile measures. In each case with a genetic disorder, parents were counseled regarding the nature and prognosis of their child’s diagnosis, as well as risk of recurrence in future pregnancies.

## Discussion

Infants with genetic disorders often exhibit limited, nonspecific phenotypes.^1,12^ Prompt genetic diagnosis may predict future manifestations and inform clinical decisions and clinical management. Our investigation demonstrated that rapidly providing genetic diagnoses altered the care of infants with CHD. We demonstrated that a multidisciplinary team of neonatologists, cardiologists, geneticists, and genetic counselors were able to identify individuals likely to benefit from testing, to return results in approximately one week, and to inform the clinical care of most infants with diagnostic results. Timely diagnoses made by rGS helped tailor of goals of care for two families. In these cases, definitive diagnoses enabled providers and families to avoid interventions which would have been intensive, costly, futile, or inconsistent with a family’s desired long-term goals. Timely genetic diagnoses also facilitated referral to subspecialists (most often ophthalmologists) and subsequent clinical diagnosis and treatment of eye disease in three cases. Eye disease is often not clinically apparent until later in childhood. Early diagnosis and correction is associated with improved vision that provides more accurate sensory input, supports early brain development, and lowers the risk of lifelong impairment.^32^

There were several patterns in the diagnoses made in this cohort by rGS. SGA birth was strongly associated with the presence of genetic disorders. Most (11 of 14) genetic disorders were unique to individuals; three cases of total anomalous pulmonary venous connection (TAPVC) were found to have cat eye syndrome (Online Mendelian Inheritance in Man Phenotype 115470) caused by mosaic supernumerary marker chromosome containing material from 22q11. The overall diagnostic heterogeneity was consistent with previous descriptions of the genetic architecture of CHD being caused by many genetic disorders.^33^ Cat eye syndrome often includes CHD, typically VSDs;^34^ however, TAPVC has recently been described in infants with cat eye syndrome.^35,36^ Genetic disorders included nine single gene (monogenic) disorders and four CNVs, highlighting the utility of rGS to diagnose both variant classes in a single test. Most (12 of 13) individuals with confirmed molecular genetic diagnoses had extracardiac anomalies. The full phenotypic spectrum of this cohort is difficult to assess as many extracardiac features, such as developmental delays, are not apparent until later in life.^18^ Family history of CHD was not associated with diagnostic rGS, and most (12 of 14) molecular diagnoses/variants were *de novo. De novo* variants are one of the most frequently recognized genetic causes of CHD.^33^

In addition to the clinical utility of a genetic diagnosis, utility was noted in non-diagnostic results. For example, the clinical team was concerned about the possibility of alveolar capillary dysplasia (ACD) in an infant with respiratory failure. Nondiagnostic rGS results in this case lowered ACD in the team’s differential and helped clarify attention on other disease processes.

There were several important limitations to this study. This study excluded individuals with known prenatal molecular genetic diagnoses. At our institution, genetic testing for structural anomalies is frequently performed prenatally.^37^ Therefore, this cohort was biased to include postnatal diagnoses, cases with prenatal care outside our medical center, and families that declined prenatal testing. Our CNICU serves as a regional referral center for complex CHD. Therefore, this cohort was also biased towards severe forms of CHD. The size of this cohort and these biases limit generalization. Finally, while we demonstrated change in clinical care using rGS, we have not yet assessed other outcomes such as length of hospitalization or economic impact, long term clinical outcomes, and psychosocial impact on parents/families and compared them to a control group.

Our model of care relied on empowering providers from multiple disciplines to work collaboratively. Neonatologists were able to nominate cases directly for rGS with genetic counselors integrated into daily rounds to identify appropriate cases and quickly conduct pretest counseling. Including genetic counselors in the cardiac neonatology team provided opportunities for reciprocal education and optimization of clinical genetic services. Finally, medical geneticists were able to focus their efforts more intensively on posttest counseling, specifically in those cases with a molecular diagnosis. This cooperative model enabled providers from each discipline to operate within their specialized skill set and led to a more efficient use of their limited time and this expensive diagnostic resource. Close involvement of geneticists and genetic counselors also demonstrates how providers from other disciplines may gain a better understanding of the application, benefits, and limitations of genome sequencing in a specific clinical context.^39^ Our model also demonstrates an equitable delivery of genomic medicine. As a regional referral center, our CNICU provides care to a broad population, disproportionately representative of underserved communities. In addition to efficiently bundling the services of highly specialized physicians, surgeons, and genetic counselors, care at our center facilitated an economy of scale that made resource-intensive rGS testing (including adequate pre- and posttest counseling) possible.

In summary, we report on implementation of rGS in infants with CHD. We found that rGS provided molecular diagnoses in 27.1% (13 of 48) of cases in approximately seven days and led to changes in clinical management in 61.5% (eight of 13) of diagnoses. In two cases rGS diagnoses helped determine goals of care and avoid intensive interventions which were futile or inconsistent with families’ goals. In three cases rGS diagnoses led to early treatment of eye disease. These findings demonstrate that clinicians should strongly consider rGS for infants presenting with CHD, especially those born SGA. Our study identified important areas for further research. We plan to evaluate the parental experience of rGS diagnostics in CHD and compare prenatal to early postnatal testing experiences. Long-term follow-up is warranted to evaluate the clinical utility of rGS to improve cardiac outcomes and to screen for and prevent extracardiac manifestations, specifically early neurodevelopmental interventions. Finally, given the extraordinary costs associated with care of infants with severe CHD (including surgery, extracorporeal membrane oxygenation, specialized nursing care, and providers from multiple disciplines), research is needed to assess the health economics of rGS for infants with CHD.

## Data Availability

All data produced in the present work are contained in the manuscript.

## Acknowledgements, Sources of Funding, and Disclosures

## Acknowledgements

none

